# Nutritional Status in Patients with Acute heart failure: A systematic review and meta-analysis with trial sequential analysis

**DOI:** 10.1101/2021.01.09.21249490

**Authors:** Jihane Belayachi, Imane Katir, Rhita Nechba Bennis, Naoufel Madani, Redouane Abouqal

**Affiliations:** Acute Medical Unit, Ibn Sina University Hospital, 10000, Rabat, Morocco; Mohammed V University in Rabat, Faculty of Medicine and pharmacy-10000, Morocco; Laboratory of Biostatistics, Clinical and Epidemiological Research, Faculty of Medicine and Pharmacy of Rabat, Morocco

**Keywords:** Acute heart failure, Malnutrition, Meta-analysis, Nutritional assessment, Prognosis

## Abstract

A critical review of the prognosis impact of malnutrition in patients admitted with acute heart failure (AHF) has never been performed. We systematically reviewed the observational epidemiology literature to determine the all-cause mortality (ACM) in undernourished patients with acute heart failure or at risk of malnutrition through a meta-analysis of observational studies.

A systemic search using PubMed, Scopus, and Web of Science was done for articles reporting an association between malnutrition and mortality in patient with acute heart failure published before December 2019. Original data from observational cohort studies in patients with acute heart failure at baseline, and with nutritional state evaluation at admission using screening, or assessment tools. The outcome of interest was mortality independent of the timeframe for follow up. The characteristics of the included study were collected. Data quality assessment using the Newcastle Ottawa Quality Assessment Scale. The hazard ratios (HRs) and corresponding 95% confidence intervals (CIs) were extracted. For the meta-analysis, a random-effects model was considered.

Heterogeneity between studies was assessed using Cochran Q statistics and I2 statistics. Subgroup analyses were used to identify the source of heterogeneity. A sensitive analysis was performed to reflect the influence of the individual data set on the pooled HR. Publication bias was detected using the Doi plot and Luis Furuya-Kanamori asymmetry index (LFK index). The influence of potential publication bias on results was explored by using the trim-and-fill procedure. To assess the risks of random errors, trial sequential analysis (TSA) was performed.

Seven studies were eligible for review and meta-analysis. There were 9053 participants and over 1536 events occurred. The prevalence of malnutrition varied from 33% to 78.8%. Mean follow-up varied between 189 and 951 days. ACM rates varied between 7% and 42.6%. Nutritional status is significantly associated with mortality in patients with AHF (Pooled HR=1.15;95%CI[1.08-1.23]). Considerable between-study heterogeneity was observed (I2=83%, P=0.001). Heterogeneity was partially explained by the different tools used to screen malnutrition risk, and follow-up durations used by the included studies. There was evidence of major publication bias regarding the risk of malnutrition-related to ACM. The obtained LFK index was 6.12 and suggests major asymmetry. The recalculated pooled HR that incorporates the hypothetical missing studies is 1.15; 95%CI (1.08-1.22). However, the accumulating number of participants and the required information size has not yet been achieved. Then, the trial sequential monitoring boundary is inconclusive.

This first meta-analysis of the association between nutritional status in patients with acute heart failure and all-cause mortality indicated that malnutrition risk in a patient with acute heart failure was associated with increased all-cause mortality. The prognosis impact of malnutrition is real despite heterogeneity in tools and cut off for defining malnutrition and mean follow up duration. This review underlines the peremptory need for multicenter studies, for uniform guidelines for assessing nutritional status, and for reporting guidelines for prognostic studies in an acute cardiovascular setting. Better nutritional practice to improve patient care is emphasized in international and national health care guidelines.

## INTRODUCTION

The incidence of heart failure (HF) has been rapidly increasing due to the ageing of society and improvement in the outcomes of cardiovascular diseases. Although evidence-based management of HF has been reported, HF still has a significant impact on the overall mortality and morbidity.[1] Patients who had been admitted for the exacerbation of HF showed 1-year mortality and readmission rates of >20%. [2] Managing patients with heart failure remains a clinical challenge, due to its underlying heterogeneity, multiple co-morbidities, and the lack of consensus on effective treatments [3] Interventions leading to early recognition of potentially modifiable risk factors could offer an alternative approach. Undernutrition is an established risk factor for adverse outcomes in patients with HF.[4]

Malnutrition mechanisms in heart failure may result from the following mechanisms: a) Gastrointestinal malabsorption due to gastrointestinal oedema or congestion and the hypoperfusion of the bowel in patients with HF are associated with poor nutrition [5]. Both hepatic congestion and gastric distention contribute to an early postprandial feeling of fullness, and they could lead to intolerance of food intake. Intestinal congestion and redistribution of blood flow away from the splanchnic circulation due to increased sympathetic activity may contribute to intestinal mucosal barrier dysfunction, which causes lipopolysaccharide translocation into the systemic circulation, and may worsen malnutrition in patients with AHF [6].

b) Impaired cardiac pump function induces a systemic response characterized by neurohormonal and inflammatory activation. Both of these are initially beneficial by inducing cytoprotective responses, but both become maladaptive over time [7]. The plasma levels of inflammatory cytokines and other chemokines seem to be elevated in direct proportion to the degradation of functional class (i.e. New York Heart Association classification) and cardiac conduct (i.e. left ventricular ejection fraction (LVEF)) [8]. Chronic HF is characterized by sustained neurohormonal activation and ultimately lead to maladaptive ventricular remodelling. Elevated catecholamine levels may lead to increased insulin resistance. This neurohormonal activation is systemic and results in insulin resistance in all tissues with potential impairment of its anabolic effects, thus potentially contributing to muscle wasting. [9]

A larger number of micronutrient deficiencies are associated with significantly shorter event-free survival. Such information is particularly important within the context of an association between sodium restriction and decreased intake of other micronutrients, including calcium, phosphate, thiamine, and folate, in addition to reduced intake of energy and carbohydrates [10]. Also, ischemia and oxidative stress may further reduce energy expenditure, resulting in cardiomyocyte injury [11].

In patients with acute heart failure, nutritional is correlated with cardiac hemodynamics, including right ventricular systolic pressure. Nutritional status was associated with E/e’ and RVSP but not LVEF, and contribute to the link of cardiac dysfunction, intestinal oedema, and cardiac cachexia [12]. Elevated RVSP correlates strongly with elevated end-diastolic pressure of the right ventricle and right atrial pressure, which may be related to intestinal congestion and, thereafter, malnutrition [7]. Intestinal congestion and redistribution of blood flow away from the splanchnic circulation due to increased sympathetic activity may contribute to intestinal mucosal barrier dysfunction, which causes lipopolysaccharide translocation into the systemic circulation, and may worsen malnutrition in patients with AHF [5].

The PICNIC study (Nutritional Intervention Program in Hospitalized Patients with Heart Failure who are Undernourished) results show that nutritional intervention in undernourished acute HF patients reduces the risk of all-cause death and the risk of readmission for worsening of HF [13].

in Emergency care; rapid nutritional intervention requires rapid recognition of nutritional status. Anthropometric parameters, such as BMI, triceps skinfold measurement and mid-arm circumference, and biochemical parameters, such as albumin, prealbumin, and cholesterol, are traditional nutritional evaluation indexes [14]. However, the use of these indexes alone cannot provide comprehensive and accurate indications of nutritional status. Numerous nutritional screening tools composed of multiple objective nutritional parameters and subjective data from case histories have been developed to enable improved nutritional status evaluations. thus, we systematically reviewed the observational epidemiology literature:

a. To determine the prevalence of undernutrition acutely admitted patient with acute heart failure
b. To assess screening tools used for acute evaluation of undernutrition
c. To evaluate the impact of survival status of patients with acute heart failure according to malnutrition risk.

Then, The current meta-analysis aimed to clarify the association between malnutrition risk in a patient with acute heart failure and all-cause mortality.

## METHODS AND MEASURES

### Literature search

The search design comprised three categories, each containing medical subject headings and keywords separated by OR. The first category defined nutritional status, the second described acute heart failure, and the third identified outcome. The exact search strategy is displayed in the *supplementary material*. Relevant studies were searched for papers published until DECEMBER 2019.

Studies were retrieved from major databases: PubMed, Scopus, and Web of Science. Articles were restricted to published literature and the English language only. No restrictions were applied regarding age, gender, or race. Duplicate papers were manually removed from the search results.

PRISMA checklist down below was used to direct the protocol of this review, and the four-phase (i.e. search, appraisal, synthesis, and analysis: SALSA) PRISMA flow diagram was used to reduce bias [15]. “This checklist has been fitted for use of protocol capitulations to Systematic Reviews from Table 2 (supplementary material) in Moher D et al.: Preferable reporting items for systematic review and meta-analysis protocols (PRISMA-P) 2015 statement.”

### Selection of papers

Two authors independently double screened all titles and abstracts of, and they excluded papers Based on pre-defined criteria. Disagreements were resolved in a consensus review.

### Inclusion and exclusion criteria

All included Studies met of the following criteria: (1) Studies presenting original data from Prospective and retrospective observational cohort studies; (2) The exposure of interest was acute heart failure at baseline, defined with validated criteria; (3) Patients with Nutritional state evaluation at admission in study population samples using screening, or assessment tools; (4) The outcome of interest was mortality independent of the timeframe for follow up. Studies were excluded if: (1) Only patients with a defined nutritional status were included (e.g., only patients with malnutrition); (2) Nutritional status evaluation using clinical or combined clinical and biological markers; (3) Follow-up was only performed in a subgroup of undernourished patients; (4) No association between the nutritional status finding of interest and mortality was described; (5) Nutritional status NOT used to evaluate prognostication.

### Data extraction

The following data elements from each study were extracted: (1) the main characteristics of the studies, including; the name of the first author, year of publication, a region of the study population, sample size; a percentage of men and mean (or median) age. (2) Diagnostic criteria of acute heart failure (3) Tools used for nutrition evaluation; and prevalence of malnutrition (including a moderate and high risk of malnutrition) (4) Mean follow up duration (by days), and estimates related to the prognostic significance of the nutritional assessment tools, HRs IC 95%. Data were independently extracted by two investigators who reached a consensus on all the items.

### Risk of bias assessment

For cohort study quality assessment, the use of the Newcastle-Ottawa Scale was recommended ^[16].^ Then, each study was evaluated on quality according to the guidelines provided by the Ottawa–Newcastle Assessment Scale (NOS) [^17,18]^. This scale grants a maximum of nine stars to each study. The maximum total score on this scale is 9. We defined studies of high quality as those that scored the maximum of 9 stars on the Newcastle-Ottawa scale. ‘‘Good’’ was defined as a total score of 7–9, ‘‘fair’’ was defined as a total score of 4–6 and ‘‘poor’’ was defined as a total score lower than 3. The discrepancy in quality assessment was discussed and resolved by the two reviewers.

### Data analysis

We used Hazard ratios as the common measure of association across studies. Eligible studies were grouped according to the tool used for nutritional status evaluation. We report the following content: The prevalence of malnutrition has identified in patients with AHF; The effect sizes (HRs), 95% confidence interval, and p values of the measures of all-cause mortality for each group.

For the meta-analysis, a random-effects model was considered [19]. Heterogeneity between studies was assessed using Cochran Q statistics and I2 statistics [20]. As suggested by Higgins et al., I2 values of 25%, 50%, and 75% were considered low, moderate, and high, respectively [21]. For P<0.10 values of the Cochran Q statistic, it was considered statistical heterogeneity, and a random-effects model was reported. Subgroup analyses were used to identify associations between outcome and relevant study characteristics: nutritional screening tools and mean follow-up duration, as a possible source of heterogeneity. Subgroup analysis was used for classified variables: Nutritional screening tools: NRI, GNRI, PNI, and CONUT, and mean follow-up duration: less than 365 days and more than 365 days.

A sensitive analysis was performed to reflect the influence of the individual data set on the pooled HR.

Publication bias was assessed using the Doi plot and Luis Furuya-Kanamori asymmetry index (LFK index). This approach has been suggested to be more robust for meta-analyses that include less than10 studies. In the presence of symmetry, one can conclude as no publication bias but in the absence of symmetry, one can expect publication bias. This publication bias was measured by the asymmetry index (LFK index). An LFK index within ±1, out of ±1 but within ±2, and > ± 2 is to mean no asymmetry, minor asymmetry and major asymmetry, respectively. [22]. The influence of potential publication bias on results was explored by using the trim-and-fill procedure [23]. *P* < 0.05 was considered statistically significant in all analyses. All analyses were conducted using Stata 14 (StataCorp, College Station, Texas). Protocol and registration; The review protocol is available in https://figshare.com/account/articles/5924662.

### Trial sequential analysis

Meta-analyses might result in type-I errors owing to an increased risk of random error when fewer patients are involved, and due to continuous significance testing when a cumulative meta-analysis is updated with new studies [24, 25]. Therefore, to assess the risks of random errors, trial sequential analysis (TSA) was performed using Stata software package (metacumbounds command), which combines information size estimation for meta-analysis (cumulated sample size of included trials) with an adjusted threshold for statistical significance in the cumulative meta-analysis. TSA was conducted with the intention to maintain an overall 5% risk of a type I error and a power of 80%. For the calculation of the required information size, an exposition effect of a 10% increased relative risk (IRR) was anticipated using average survival event proportion calculated from the actual meta-analyses. [26]

## RESULTS

### Search results

As mentioned previously, two authors independently double screened all titles and abstracts of, and they excluded papers based on pre-defined criteria. Disagreements were resolved in a consensus review. Of the 583 articles retrieved, 23 completed articles were reviewed, and 7 studies met all inclusion criteria. The total number of patients included in the studies ranged between and 5265 with 9053 cases included. The mean age exceeded 75 years. There were five prospective cohort studies and two retrospective cohort studies. An overview of the selection procedure is shown in the flowchart in **Figure 1**. Methodological aspects of the included studies, the study population, completeness and duration of follow-up, definition of prognostic variables, and outcome were clearly described in most studies.

**Figure 1:**
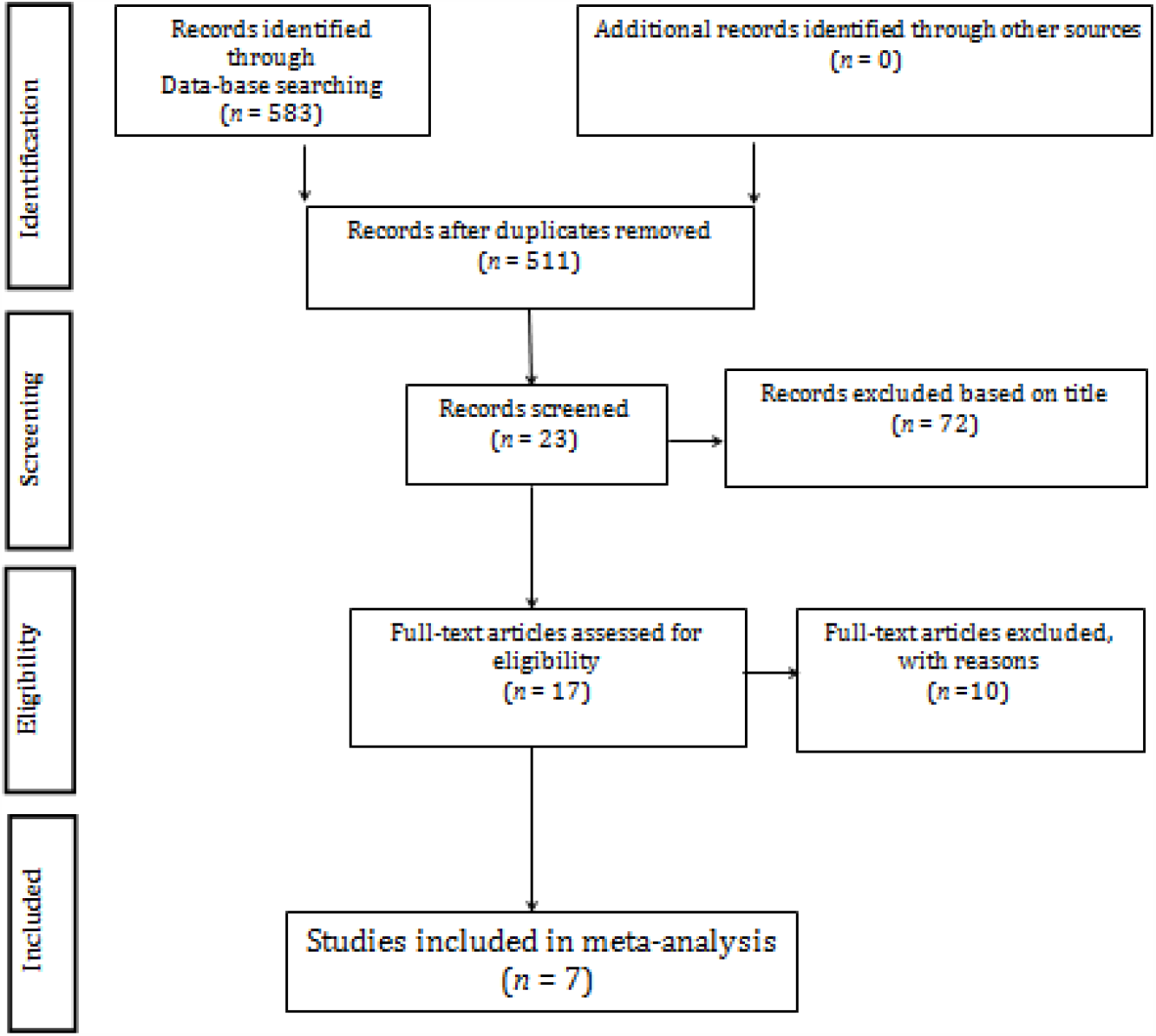
Flow chart of study selection process.

### Characteristics of studies

Studies showed good quality assessment; detailed quality assessment of the seven studies included in the meta-analysis are presented in **Table 3** (supplementary material).

The characteristics of the studies included seven prospective studies and two retrospective cohort studies. There were 9053 participants and over 1536 events occurred, including cardiovascular mortality and1386 all-cause mortality. The cohorts were from four different countries: Japan mostly, Taiwan; Korea and the United Kingdom. Study lengths ranged from October 2003 till June 2016. Multiple adjustments criteria were included in the review studies and the most frequent confounders that were adjusted were: Age; Gender; BMI; Renal function; Total bilirubin, haemoglobin, and sodium; History of heart failure; History of hypertension, diabetes, dyslipidemia, smoking and drugs’ prescription. Studies assessing prognosis impact of undernutrition in a patient with acute heart failure were summarized in presented in **Table 1**

**Table 1:** **Summary of studies assessing prognosis impact of undernutrition in patient with acute heart failure and included in the analysis**

| Author | Country | Sample | Mean Age (years) | Male gender (%) | AHF | Prevalence of malnutrition (according to screening tool) | Mean Duration of follow-up (days) | ACM |
| --- | --- | --- | --- | --- | --- | --- | --- | --- |
| Iwakami 2016 [32] | Japan | 635 | 75 ± 12 | 62 | Framingham criteria [27]<br>De novo HF; Decompensation of chronic HF; HFrEF and HFpEF. | CONUT 77.6% | 324 | 10% |
| Honda Y 2016 [35] | Japan | 490 | 79±7 | 59 | Framingham criteria [27]<br>De Novo HF; Decompensation of chronic HF; HFrEF and HFpEF. | GNRI 33% | 189 | 11.2% |
| Cheng TL 2017 [12] | Taiwan | 1673 | 76±13 | 68 | American Society of Echocardiography [28]<br>De Novo HF; HFrEF and HFpEF. | PNI 66.5% | 951 | 44.3% |
| Shirakabe A 2017 [34] | Japan | 458 | 76 (67-82) | 66 | ESC [29]<br>New-onset HF; Decompensation of chronic HF. | CONUT 71.9%<br>PNI 27.7% | 365 | 7% |
| Yeong Cho 2018 [33] | Korea | 5265 | 68.4 ±14.5 | 52.8 | ESC [29]<br>De novo HF; Decompensation of chronic HF/ ADHF; Hypertensive HF; Pulmonary edema; Cardiogenic shock; Right HF. | NRI 67.3%<br>PNI 42%<br>GNRI 46% | 365 | 5.1% |
| Sze S 2017 [36] | UK | 265 | 80 | 62 | ND Primary diagnosis of HF secondary to LVSD* | CONUT 46%<br>GNRI 42% | 598 | 42.6% |
| Ouchi 2017 [37] | Japan | 267 | 73(64-82) | 53.7 | Framingham criteria [27] | GNRI 41.9% | 720(22±14.2 months) | 23.2% |

### Definition of AHF

Many definitions were suggested in the reviewed studies based on different societies. Two of them did not define the AHF and only referred to the symptomatic characteristics of heart failure. Three studies adapted the Framingham criteria [27]. Cheng YL and Al. went with the American Society of Echocardiography’s definition [28]. And finally, two studies deployed the interpretation of the European Society of Cardiology (ESC) [29]. Jae Yeong Cho and Shirakabe A referred to the European Society of Cardiology that defined the HF as a clinical syndrome characterized by typical symptoms (such as breathlessness, ankle swelling and fatigue) that may be accompanied by signs (such as high oedema, high jugular venous pressure, and pulmonary hisses) caused by a structural and/or functional cardiac abnormality, resulting in reduced cardiac output and/or elevated intracardiac pressures at rest or during stress [30,31]. ***Table 1*** *summarized* the definitions used in the different studies reviewed.

### Modalities of screening tools used in AHF patients

#### Instruments

##### Participants characteristics

The population studied in the review articles had a wide sample ranging between 265 patients to 5265 with a total of 9053 cases of patients. The prevalence of malnutrition and the risk of malnutrition varied among the systematically retrieved studies from 33% to78.8%. However, this range of prevalence’s included only studies with pre-defined cut-off. However, different cut off was used for the same tool depending on studies (Table 4 in supplementary material). The studies included investigated in-hospital and post-discharge all-cause mortality. Mean follow-up varied between 189 and 951 days. All-cause-mortality and rates vary between 7% and 42.6%. **Table 1** summarized study characteristics.

### Prognosis impact of malnutrition in patients with AHF: Results of individual studies

- Cheng and al [12]. evaluated the model performance of PNI compared to each of its components alone and revealed more prognostic predictive power. In this
- study, the authors found that a lower PNI was associated with increased mortality after adjusting for other variables.
- Iwakami and al [32]. showed that per point increase in the CONUT score was associated with an increased risk of all-cause death, even after full adjustment by major confounders.
- Yeong Cho and al [33]. found that NRI was a predictor of composite endpoints of all-cause mortality and readmission rates with episodes of ADHF.
- Shirakabe and al [34] applied simultaneous PNI and CONUT to predict mortality. PNI exhibited a good balance between sensitivity and specificity for predicting in-hospital mortality.
- Honda and al [35] showed that malnutrition evaluated by lower GNRI score on admission was independently associated with increased mortality in patients with AHF.
- Both Sze [***36***] and Cheng demonstrated that nutrition status, based on PNI in patients with AHF, was independently associated with both short- and long-term all-cause and cardiovascular mortality.
- Ouchi et al [37] showed that all-cause death was higher in patients with high GNRI than in those with low or GNRI.

### Prognosis impact of malnutrition in patients with AHF: Meta-Analysis

Nutritional status is significantly associated within patients with risk of malnutrition (the pooled HR=1.15 [1.08 - 1.23]) with statistical heterogeneity among studies (I2 = 83%, P = 0.001). **Figure 2** presented the Forest plot for risk of ACM associated with the risk of malnutrition in patients with AHF.

**Figure 2:**
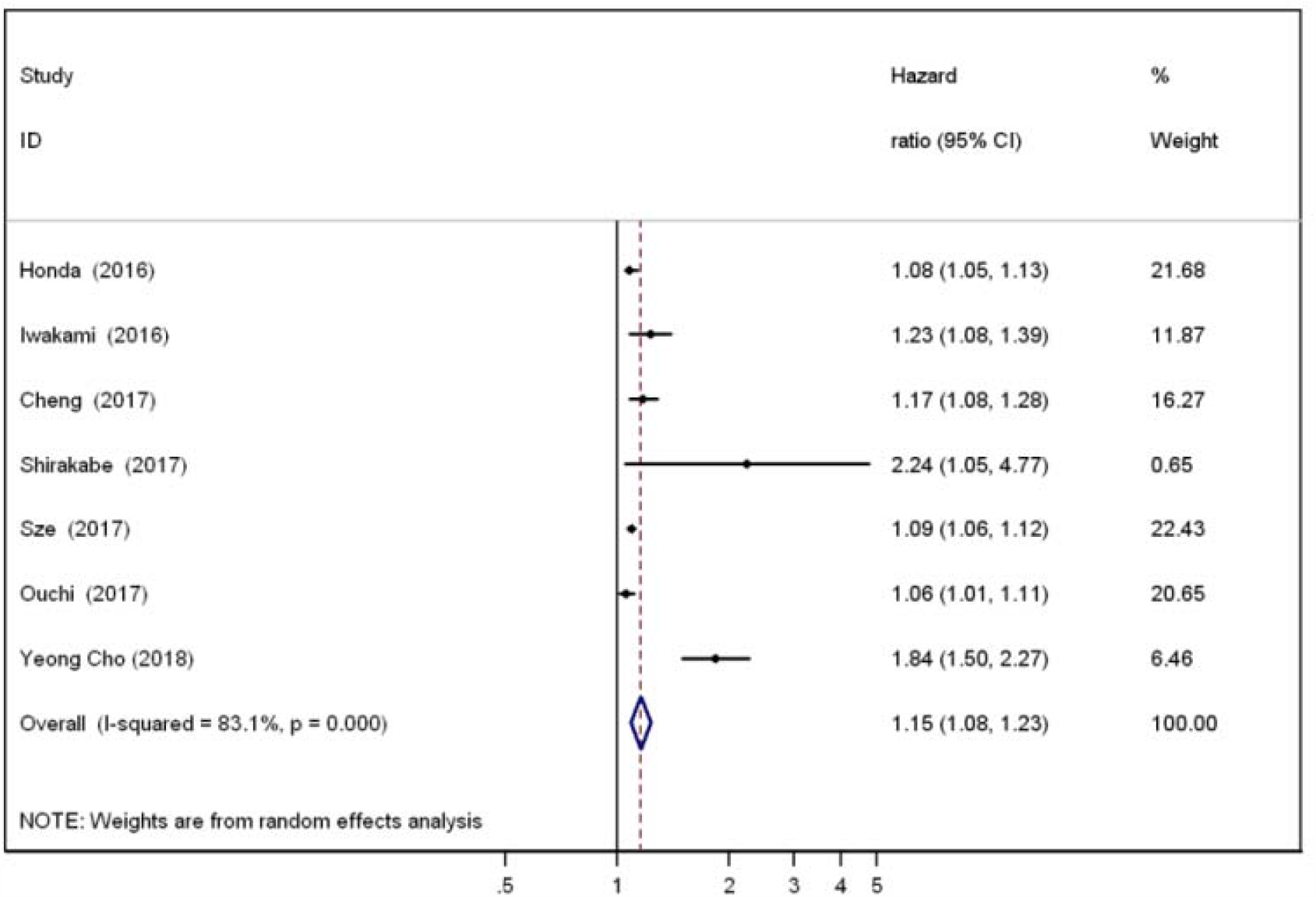
Forest plot for risk of ACM associated with risk of malnutrition in patients with AHF.

### Sources of heterogeneity

Subgroup analyses were used to identify associations between outcome and nutritional screening tools and the mean follow-up duration as possible sources of heterogeneity.

### Nutritional screening tool

There was no longer any evidence of significant heterogeneity in studies using GNRI (I2 = 0%), however studies using PNI (2 studies) and CONUT (2 studies) explain respectively (I2 = 58.6% and I2 = 57.2%) of heterogeneity between studies. Association between ACM and malnutrition risk in each tool showed significant association except for studies using CONUT: The polled HR =1.47 (0.86 to 2.51) indicating no association between ACM and malnutrition risk assessed by CONUT. **Figure 3** presented a forest plot of the association between mortality and malnutrition by the subgroup of screening tools.

**Figure 3:**
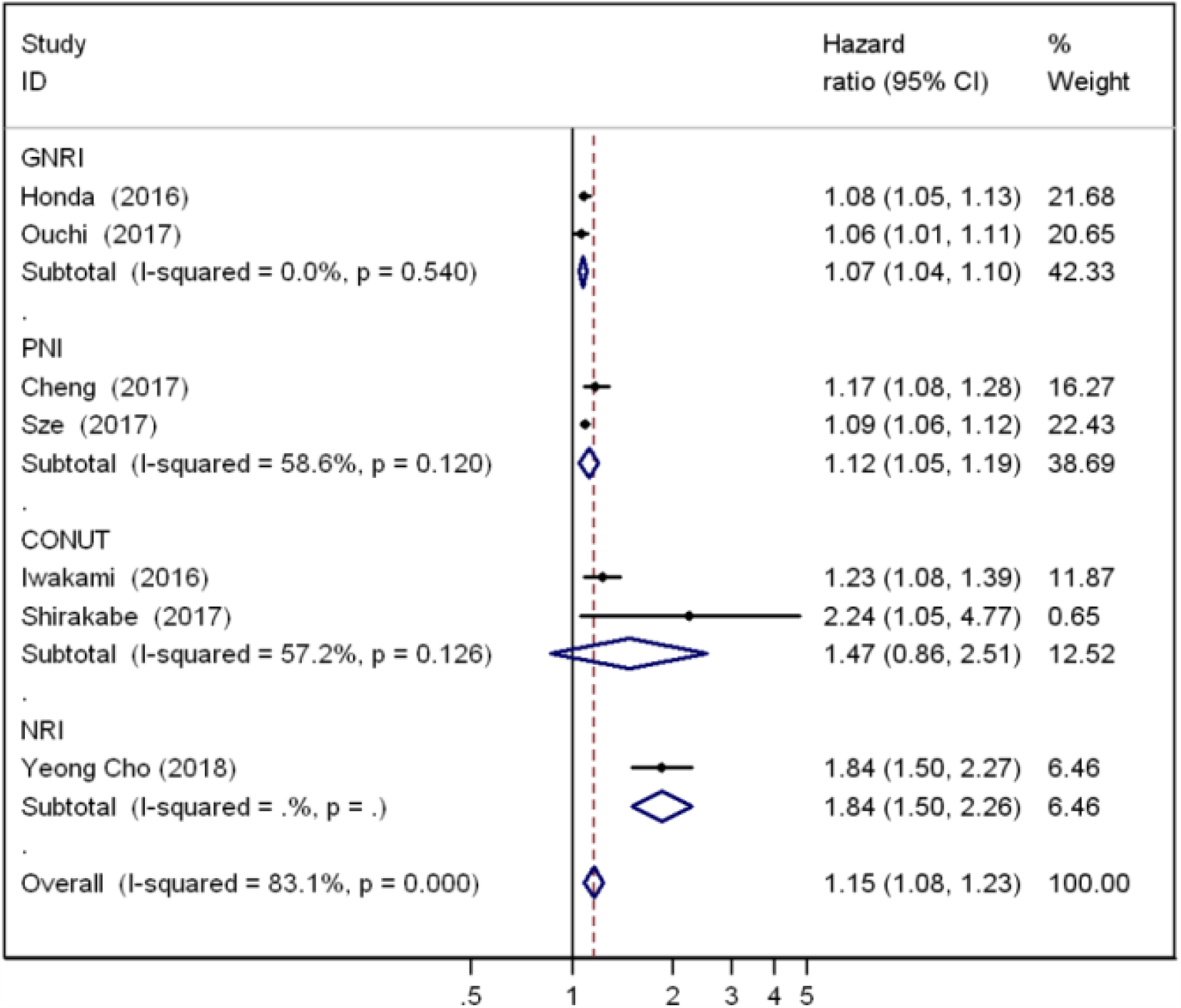
Forest plot of the association between mortality and malnutrition by the subgroup of screening tools.

### Mean follow-up duration

Heterogeneity between studies was mainly explained by mean follow up duration less than 365 days in 3 studies (I2 = 83%; p= 0.002). There was no longer any evidence of significant heterogeneity in 4 studies with 365 days means to follow up duration and more (I2 = 27%; p= 0.24). Forest plot of the association between mortality and malnutrition by the subgroup of the mean follow-up duration is presented in the supplementary material Figure 1.

### Publication Bias

There was evidence of major publication bias regarding the risk of malnutrition-related ACM as indicated by the LFK index. The obtained LFK index was 6.12 and suggests major asymmetry. Doi plot based on all-cause mortality in patient with AHF with malnutrition risk was presented in **figure 4**. The ‘‘trim and fill’’ procedure found one possible ‘‘missing’’ study. The recalculated pooled Hazard ratio that incorporates the hypothetical missing studies is 1.15 (1.08-1.22).

**Figure 4:**
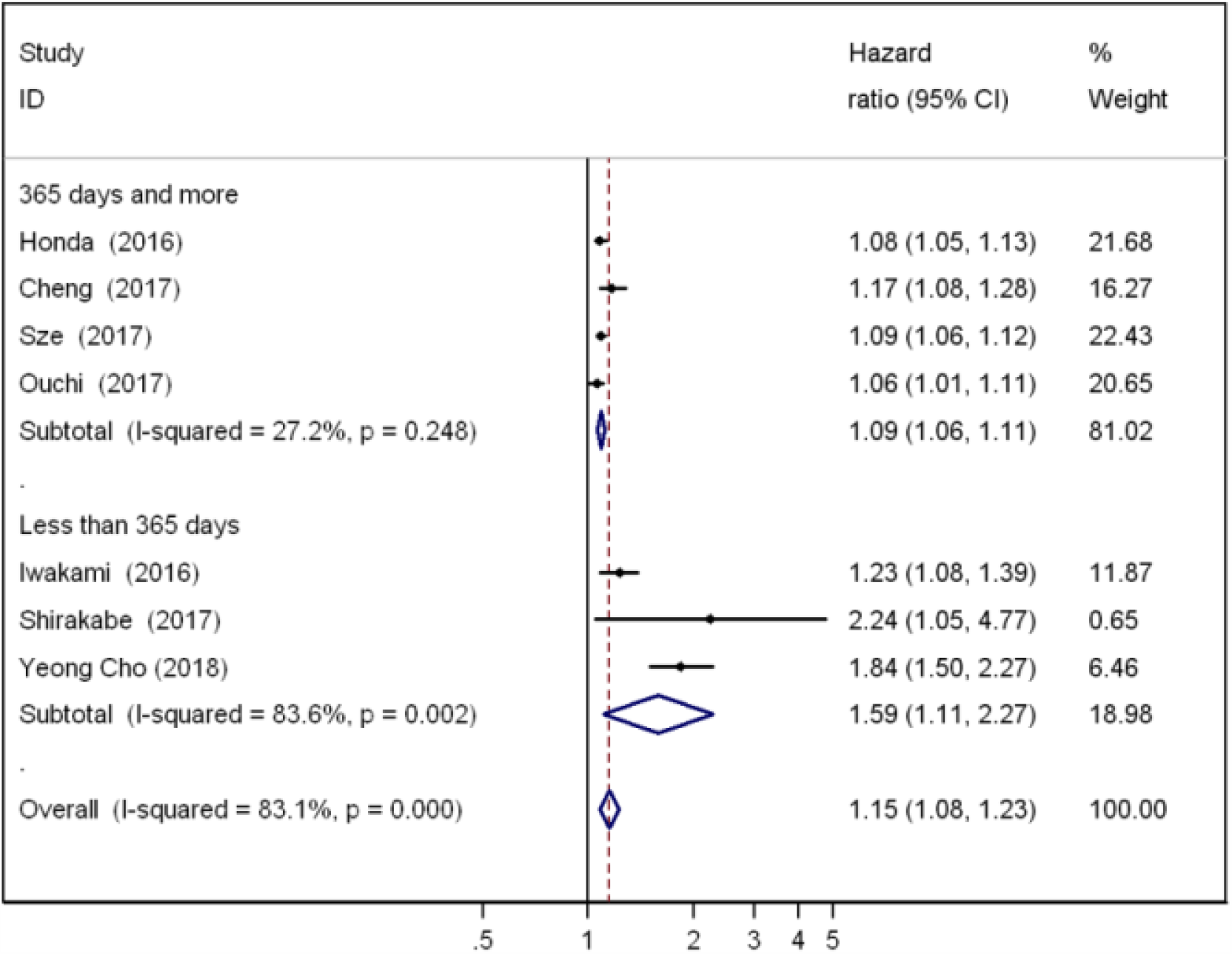
Forest plot of the association between mortality and malnutrition by the subgroup of the mean follow-up duration.

### Trial Sequential Analysis

The required information size to detect the 10% increased relative risk found in the random-effects model meta-analysis is calculated to 9053 participants. The cumulative Z-curve surpasses the traditional boundary and the trial sequential monitoring boundaries for statistical significance. However, the accumulating number of participants and the required information size has not yet been achieved. Then, the trial sequential monitoring boundary is inconclusive. Trial Sequential Analysis of survival status according to nutritional status in acute heart failure patient is presented in **figure 5**.

**Figure 5:**
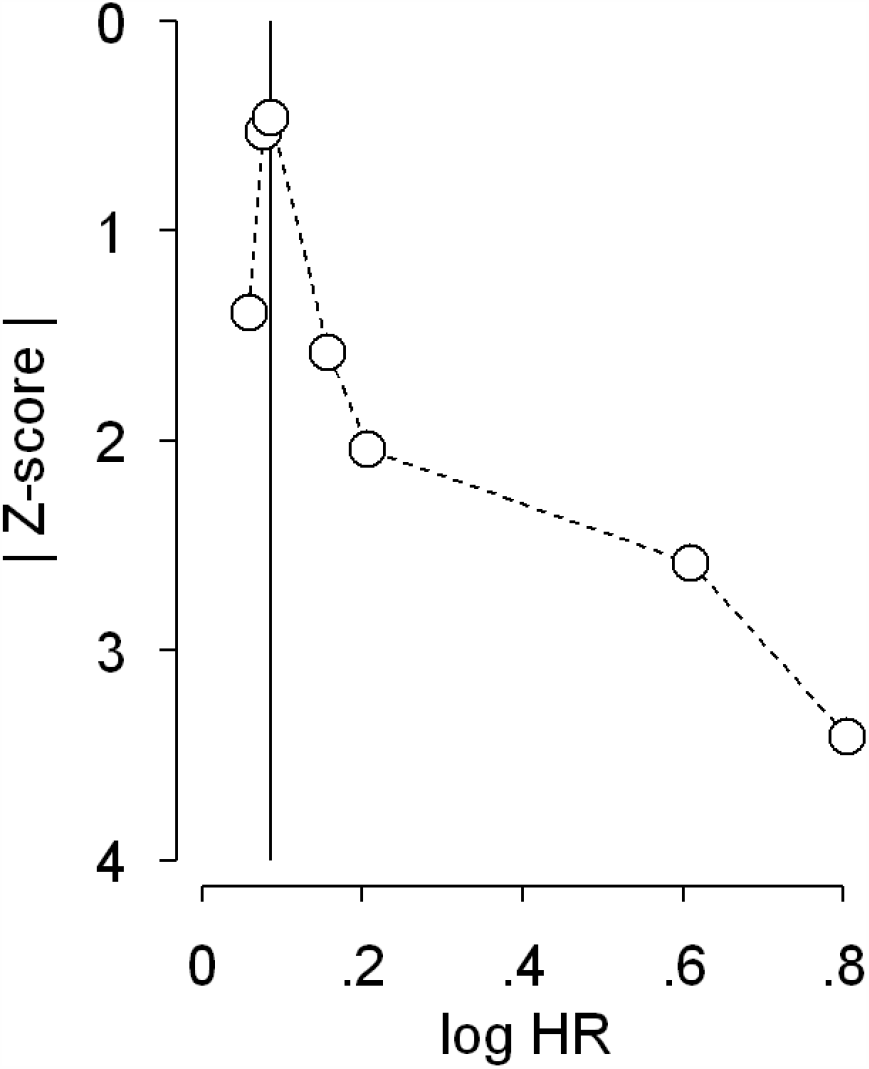
Doi plot based on all-cause mortality in patient with AHF with malnutrition risk. The vertical line on the horizontal (x) axis represents the effect size (ES) with the lowest absolute z score, dividing the plot into two regions with the same areas. Visualisation of the plot suggests major asymmetry and thus small-study effects such as publication bias. The obtained Luis Furuya Kanamori index of 6.12 also suggests major assymmetry.

**Figure 6.**
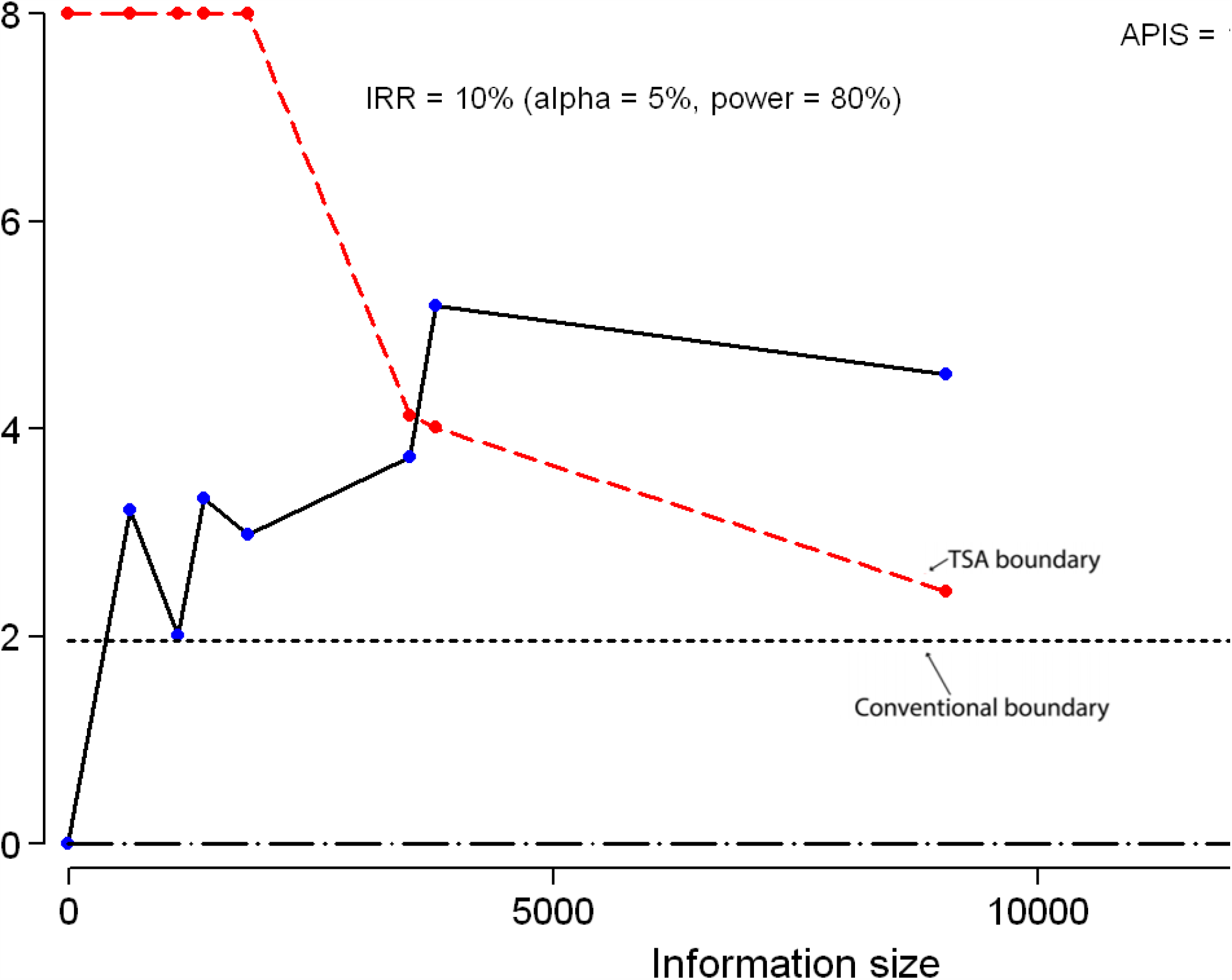
Trial Sequential Analysis of survival status according to nutritional status in acute heart failure patient. Trial sequential monitoring boundary, is inconclusive because the accumulating number of participants and the required information size has not yet been achieved. the evidence was not conclusive for the outcome. The Z-value is the test statistic and |*Z*|⍰=⍰ 1.96 corresponds to a *P*⍰ =⍰ 0.05; the higher the *Z*-value, the lower the *P*-value. The required information size to detect the 10% increased relative risk found in the random-effects model meta-analysis is calculated to 9053 participants. The cumulative *Z*-curve (black full line with indications of each trial) surpasses the traditional boundary and the trial sequential monitoring boundaries (RED curves) for statistical significance have been surpassed in the TSA. **APIS: A priori information size**

## Discussion

We conducted the first comprehensive analysis of the association between Nutritional Status in Patients with Acute heart failure and ACM through a meta-analysis of observational studies.

The meta-analysis showed that malnutrition risk in a patient with acute heart failure was associated with increased all-cause mortality. Studies investigated in survival outcome with follow up between 189 days and 951 days and mean follow up duration explain heterogeneity between studies. Since study with longer follow-up is less heterogenic.

The prevalence of malnutrition in a patient admitted with acute heart failure varied among the studies from 33% and 78.8%. Estimates of the incidence of HF-associated malnutrition vary according to the population studied, and the criteria used to define malnutrition. We reported a great variety of tools used to evaluate nutritional status and on cut off for defining malnutrition which, reasons for the differences in the prevalence of undernutrition include the acquisition of data using different nutritional screening tools. We reported a great variety of tools used to evaluate nutritional status, and one cut off for defining malnutrition which creates a discrepancy in prevalence between studies, and also mortality rates at a different time follows up. The tools used in this review were based on serological markers, which raise the question concerning the use of these nutritional tools in an acute context of inflation and inflammation.

Diagnosis of acute heart failure was based on the clinical definition in an acute situation. However, the heart failure spectrum and aetiology of heart failure are not clearly defined. In this review, this gap can explain variation in malnutrition prevalence and mortality rates Heterogeneity of heart failure is manifested by the varying profiles of ventricular remodelling among patients with heart failure. Each heart failure phenotype is the result of a patient-specific trajectory wherein the heart remodels towards concentric hypertrophy, eccentric hypertrophy, or a combination of both. The way of entry and the subsequent path of the trajectory depends on the patient’s risk factors, comorbidities, and disease modifiers. Risk factors are disease entities that always precede the development of heart failure and are associated with increased heart failure incidence. Comorbidities may precede or develop after heart failure and usually coexist with heart failure in groups of two or more (multi-morbidity). Modifiers are specific patient characteristics that contribute to the development of the entry phenotype and heart failure progression. Since the authors adjusted the HR to preexisting heart failure, but not spectrum and aetiology heart failure [38].

As mentioned earlier, nutritional screening is the first step towards differencing between malnourished and non-malnourished patients. However, there is no international consensus on a single ‘best tool’. The use of different tools in different studies isn’t compatible with a comparison between studies and does not allow for the drawing of conclusions on defining the ‘best tool’ for a certain patient population, age group or setting.

NRI has little advantage over serum albumin in terms of nutritional assessment in AHF [39]. Nevertheless, it is considered that NRI may not be useful for patients aged ≥ 65 years because of difficulty in determining their usual body weight at home. From this point of view, Bouillanne and Al. modified the NRI score and developed GNRI using ideal body weight instead of usual body weight [40].

GNRI is hypothesized to be an appropriate tool for evaluating nutritional status in the decompensated phase of HF because increased extracellular fluid volume decreases serum albumin, whereas it increases actual body weight. However, GNRI was considered the weakest predictor of mortality, perhaps because GNRI includes weight loss. Weight loss is unreliable in patients with HF because of the influence of oedema and the use of diuretics [41].

CONUT was originally proposed by Ignacio de Ulíbarri and Al. as a screening tool for malnutrition in hospitalized patients and the score consists of three indices: Serum albumin; Total cholesterol; and lymphocyte count.

In this meta-analysis, The polled HR indicating no association between ACM and malnutrition risk assessed by CONUT. The CONUT score is thus appropriate for evaluating diverse aspects of the complex mechanism of malnutrition in HF because each of the three components reflects different aspects of malnutrition. PNI is similar to the CONUT score but does not include cholesterol, which might be more appropriate in patients with HF as a significant proportion takes statins which cause lower cholesterol levels irrespective of nutritional status. Cheng YL and Al. Investigated for the first time the clinical significance of PNI in patients hospitalized for AHF and demonstrated that PNI revealed more prognostic predictive power than its components alone and that PNI in patients with AHF, was independently associated with both short-term and long-term total and cardiovascular mortality. Compared to other screening tools, two studies found that PNI has the best model performance [20, 42].

The prognosis impact of malnutrition is real despite heterogeneity in tools and cut off for defining malnutrition, in the heart failure spectrum phenotype and mean follow up duration. This review underlines the peremptory need for multicenter studies, for uniform guidelines for assessing nutritional status, and for reporting guidelines for prognostic studies in an acute cardiovascular setting. Overall, malnutrition is often observed in patients with acute heart failure and is associated with adverse clinical outcomes. Better nutritional practice to improve patient care is emphasized in international and national health care guidelines.

This first meta-analysis of the association between Nutritional Status in Patients with Acute heart failure and all-cause mortality indicated that malnutrition risk in a patient with acute heart failure was associated with increased all-cause mortality. The prognosis impact of malnutrition is real despite heterogeneity in tools and cut off for defining malnutrition, in the heart failure spectrum phenotype and mean follow up duration. This review underlines the peremptory need for multicenter studies, for uniform guidelines for assessing nutritional status, and for reporting guidelines for prognostic studies in an acute cardiovascular setting. Overall, malnutrition is often observed in patients with acute heart failure and is associated with adverse clinical outcomes. Better nutritional practice to improve patient care is emphasized in international and national health care guidelines. Contemporaneousness concern in the evaluation of all aspects of healthcare using science-based methods speculates that nutrition screening programs be utterly evaluated [43].

## Data Availability

https://figshare.com/account/articles/5924662.

## References

1. Conrad N, Judge A, Tran J, Mohseni H, Hedgecott D, Crespillo A.P, et al (2018).Temporal trends and patterns in heart failure incidence: a population-based study of 4 million individuals. Lancet 391:572–580.

2. Taylor C.J, Ordonez-Mena J.M, Roalfe A.K, Lay-Flurrie S, Jones N.R, Marshall T, et al (2019). Trends in survival after a diagnosis of heart failure in the United Kingdom 2000–2017: population based cohort study. BMJ 364: 223

3. Redfield MM (2016). Heart failure with preserved ejection fraction. N Engl J Med 375: 1868– 1877.

4. Narumi T, Arimoto T, Funayama A, Kadowaki S, Otaki Y, Nishiyama S, et al (2013). Prognostic importance of objective nutritional indexes in patients with chronic heart failure. J Cardiol 62: 307–313.

5. Fernando G. Romeiro, Katashi Okoshi, Leonardo A. M. Zornoff, Marina P. Okoshi (2012). Gastrointestinal changes associated to heart failure. Arq. Bras. Cardiol 98:3.

6. Valentová M, Haehling S von, Bauditz J, Doehner W, Ebner N, Bekfani T, et al (2016). Intestinal congestion and right ventricular dysfunction: a link with appetite loss, inflammation, and cachexia in chronic heart failure. Eur Heart J: 37:1684⍰91.

7. Justin Hartupee, Douglas L. Mann. Neurohormonal activation in heart failure with reduced ejection fraction (2017). Nat Rev Cardiol 14(1): 30–38.

8. Mann DL. (2015) Innate immunity and the failing heart: the cytokine hypothesis revisited. Circ Res 116(7):1254⍰68.

9. Anker SD, Sharma R. (2002) The syndrome of cardiac cachexia. Int J Cardiol 85(1)

10. Alberto Miján-de-la-Torre.(2009) Recent insights on chronic heart failure, cachexia and nutrition. Curr Opin Clin Nutr Metab Care 12(3):251.

11. Gino A. Kurian, Rashmi Rajagopal,Srinivasan Vedantham, and Mohanraj Rajesh. (2016) The Role of Oxidative Stress in Myocardial Ischemia and Reperfusion Injury and Remodeling: Revisited Oxid Med Cell Longev 1656450.

12. Cheng Y-L, Sung S-H, Cheng H-M, Hsu P-F, Guo C-Y, Yu W-C, et al (2017). Prognostic Nutritional Index and the Risk of Mortality in Patients With Acute Heart Failure. J Am Heart Assoc 25: 6(6).

13. Bonilla-Palomas L, Gámez-López A.L, Castillo-Domínguez J.C, Moreno-Conde López-Ibáñez M, Alhambra-Expósito R, et al (2016). Nutritional intervention in malnourished hospitalized patients with heart failure. Arch Med Res 47: 535–540.

14. Kondrup J, Allison SP, Elia M, Vellas B, Plauth M,(2003) Educational and Clinical Practice Committee, European Society of Parenteral and Enteral Nutrition (ESPEN). ESPEN guidelines for nutrition screening. Clin Nutr Edinb Scotl 22(4): 415⍰21.

15. Liberati A, Altman DG, Tetzlaff J, Mulrow C, Gøtzsche PC, Ioannidis JPA, et al (2009). The PRISMA statement for reporting systematic reviews and meta-analyses of studies that evaluate health care interventions: explanation and elaboration. PLoS Med 6(7)

16. Zeng X, Zhang Y, Kwong JSW, Zhang C, Li S, Sun F, et al (2015). The methodological quality assessment tools for preclinical and clinical studies, systematic review and meta-analysis, and clinical practice guideline: a systematic review. J Evid-Based Med 8(1):2⍰10.

17. Ottawa Hospital Research Institute. http://www.ohri.ca/programs/clinical_epidemiology/oxford.asp (available 20 déc 2019]

18. Stang A (2010). Critical evaluation of the Newcastle-Ottawa scale for the assessment of the quality of nonrandomized studies in meta-analyses. Eur J Epidemiol 25(9):603⍰5.

19. DerSimonian R, Laird N (1986). Meta-analysis in clinical trials Control Clin Trials 7(3):177–88

20. Julian P T Higgins, Simon G Thompson, Jonathan J Deeks, Douglas G Altman (2003). Measuring inconsistency in meta-analyses. – BMJ 327:557–60.

21. Julian PT. Higgins; Simon G. Thompson. Quantifying heterogeneity in a meta-analysis - Statistics in Medicine 2002.

22. Furuya-Kanamori L, Barendregt JJ, Doi SAR A (2018) new improved graphical and quantitative method for detecting bias in meta-analysis. Int J Evid Based Health 16: 195– 203.

23. Duval S, Tweedie R. (2000) Trim and fill: A simple funnel-plot-based method of testing and adjusting for publication bias in meta-analysis. Biometrics 56:455–63. 10.1111/j.0006-341X.2000.00455.x

24. Wetterslev J, Thorlund K, Brok J, Gluud C. (2008) Trial sequential analysis may establish when firm evidence is reached in cumulative meta-analysis. J Clin Epidemiol. 61:64–75.

25. Brok J, Thorlund K, Wetterslev J, Gluud C. (2009). Apparently conclusive meta-analyses may be inconclusive–trial sequential analysis adjustment of random error risk due to repetitive testing of accumulating data in apparently conclusive neonatal meta-analyses. Int J Epidemiol. 38:287–98.

26. Branko Miladinovic, Iztok Hozo, Benjamin Djulbegovic (2013). Trial Sequential Boundaries for Cumulative Meta-Analyses. The Stata Journal 13(1): 77–91

27. William Kannel. Framingham Heart Failure Diagnostic Criteria https://www.mdcalc.com/framingham-heart-failure-diagnostic-criteria.

28. John Gorcsan, Theodore Abraham, Deborah A Agler, Jeroen J Bax, Genevieve Derumeaux, Richard A Grimm, et al (2008). Echocardiography for Cardiac Resynchronization Therapy: Recommendations for Performance and Reporting–A Report from the American Society of Echocardiography Dyssynchrony Writing Group Endorsed by the Heart Rhythm Society J Am Soc Echocardiogr 21(3):191–213.

29. McMurray JJV, Adamopoulos S, Anker SD, Auricchio A, Böhm M, Dickstein K, et al (2012). ESC guidelines for the diagnosis and treatment of acute and chronic heart failure 2012: The Task Force for the Diagnosis and Treatment of Acute and Chronic Heart Failure 2012 of the European Society of Cardiology. Developed in collaboration with the Heart Failure Association (HFA) of the ESC. Eur J Heart Fail 14(8):803⍰69.

30. Nussbaumerová B, Rosolová H (2018). Diagnosis of heart failure: the new classification of heart failure. Vnitr Lek. Fall 64(9):847⍰51.

31. Piotr Ponikowski, Adriaan A Voors, Stefan D Anker, Héctor Bueno, John G F Cleland, Andrew J S Coats, Volkmar Falk et al. ESC Guidelines for the diagnosis and treatment of acute and chronic heart failure. European Heart Journal 37(27):2129–2200

32. Iwakami N, Nagai T, Furukawa TA, Sugano Y, Honda S, Okada A, et al (2017) Prognostic value of malnutrition assessed by Controlling Nutritional Status score for long-term mortality in patients with acute heart failure. Int J Cardiol 230:529⍰36.

33. Cho JY, Kim KH, Cho H-J, Lee H-Y, Choi J-O, Jeon E-S, et al (2018). Nutritional risk index as a predictor of mortality in acutely decompensated heart failure. PLOS ONE 13(12):e0209088.

34. Shirakabe A, Noritake Hata, Nobuaki Kobayashi, Hirotake Okazaki, Masato Matsushita, Yusaku Shibata, Suguru Nishigoori, Saori Uchiyama, Kuniya Asai, Wataru Shimizu (2018). The prognostic impact of malnutrition in patients with severely decompensated acute heart failure, as assessed using the Prognostic Nutritional Index (PNI) and Controlling Nutritional Status (CONUT) score. Heart Vessels 33(2):134–144.

35. Honda Y, Nagai T, Iwakami N, Sugano Y, Honda S, Okada A, et al (2016). Usefulness of Geriatric Nutritional Risk Index for Assessing Nutritional Status and Its Prognostic Impact in Patients Aged ≥65 Years With Acute Heart Failure. Am J Cardiol 118(4):550⍰5.

36. Sze S, Zhang J, Pellicori P, Morgan D, Hoye A, Clark AL (2017). Prognostic value of simple frailty and malnutrition screening tools in patients with acute heart failure due to left ventricular systolic dysfunction. Clin Res Cardiol 106(7):533–541.

37. Ouchi S, Miyazaki T, Shimada K, Sugita Y, Shimizu M, Murata A, et al (2017). Low Docosahexaenoic Acid, Dihomo-Gamma-Linolenic Acid, and Arachidonic Acid Levels Associated with Long-Term Mortality in Patients with Acute Decompensated Heart Failure in Different Nutritional Statuses. Nutrients 9(9).

38. Filippos Triposkiadis, Javed Butler, Francois M Abboud, Paul W Armstrong, Stamatis Adamopoulos, John J Atherton et al (2019). The continuous heart failure spectrum: moving beyond an ejection fraction classification. Eur Heart J 1;40(26):2155–2163..

39. Buzby GP, Williford WO, Peterson OL, Crosby LO, Page CP, Reinhardt GF, et al (1988). A randomized clinical trial of total parenteral nutrition in malnourished surgical patients: the rationale and impact of previous clinical trials and pilot study on protocol design. Am J Clin Nutr 47:357⍰65

40. Bouillanne O, Morineau G, Dupont C, Coulombel I, Vincent J-P, Nicolis I, et al (2005). Geriatric Nutritional Risk Index: a new index for evaluating at-risk elderly medical patients. Am J Clin Nutr 82(4):777⍰83.

41. Wada H, Dohi T, Miyauchi K, Doi S, Naito R, Konishi H, et al (2017). Prognostic Impact of the Geriatric Nutritional Risk Index on Long-Term Outcomes in Patients Who Underwent Percutaneous Coronary Intervention. Am J Cardiol 01.

42. Kondrup J, Allison SP, Elia M, Vellas B, Plauth M, (2003). Educational and Clinical Practice Committee, European Society of Parenteral and Enteral Nutrition (ESPEN). ESPEN guidelines for nutrition screening 2002. Clin Nutr Edinb Scotl 22.

43. Charney P (2008). Nutrition screening vs nutrition assessment: how do they differ? Nutr Clin Pract Off Publ Am Soc Parenter Enter Nutr 23(4):366⍰72.

