## appendix for "Nutritional Status in Patients with Acute heart failure: A systematic review and meta-analysis with trial sequential analysis"

Table 1: Search strategy of the review

|  | **Category 1** | A  N  D | **Category 2** | A  N  D | **Category 3** |
| --- | --- | --- | --- | --- | --- |
| **M**  **E**  **S**  **H** | - Acute heart failure - Acute decompensated heart failure - Congestive heart failure |  | - Nutritional status - Malnutrition - Undernutrition - Nutritional risk - Nutritional assessment - Nutritional screening |  | "Disease-Free Survival"  "Follow-Up Studies"  "Kaplan-Meier Estimate"  "Multivariable Analysis"  "Predictive Value of Tests"  "Prognosis"  "Prospective Studies"  "Proportional Hazards Models"  "Risk Assessment"  "Survival Analysis" |
| **T**  **E**  **X**  **T** | - Primary diagnosis of HF - The first episode of rapid onset of heart failure - Worsening symptoms of heart failure - Worsening signs of heart failure - Novo HF - New-onset HF, - Decompensation of chronic HF. - New or worsening HF,AHF; ADHF; HF; AHFS |  | - Dysnutritional state - Risk of malnutrition - Risk for undernutrition |  | Clinical outcome  Cox proportional  Follow-up studies  Hazard ratio  Incremental predictor  Incremental prognostic value  Kaplan-Meier  Multivariable analysis  Prognostic value  Prognosis  Prognostic stratification  Risk stratification  Survival analysis |

Table 2: The PRISMA-check list for quality assessment.

| **Section/topic** | **#** | **Checklist item** | **Information reported** | | **Line number(s)** |
| --- | --- | --- | --- | --- | --- |
|  |  |  | **Yes** | **No** |  |
| **ADMINISTRATIVE INFORMATION** | | | | | |
| **Title** | | | | | |
| Identification | 1a | Identify the report as a protocol of a systematic review | ☒ | ☐ |  |
| Update | 1b | If the protocol is for an update of a previous systematic review, identify as such | ☐ | ☒ |  |
| **Registration** | 2 | If registered, provide the name of the registry and registration number in the Abstract | ☒ | ☐ |  |
| **Authors** | | | | | |
| Contact | 3a | Provide name, institutional affiliation, and e-mail address of all protocol authors; provide physical mailing address of corresponding author | ☒ | ☐ |  |
| Contributions | 3b | Describe contributions of protocol authors and identify the guarantor of the review | ☒ | ☐ |  |
| **Amendments** | 4 | If the protocol represents an amendment of a previously completed or published protocol, identify as such and list changes; otherwise, state plan for documenting important protocol amendments | ☐ | ☒ |  |
| **Support** | | | | | |
| Sources | 5a | Indicate sources of financial or other support for the review | ☐ | ☒ |  |
| Sponsor | 5b | Provide name for the review funder and/or sponsor | ☐ | ☒ |  |
| Role of sponsor/funder | 5c | Describe roles of funder(s), sponsor(s), and/or institution(s), if any, in developing the protocol | ☐ | ☒ |  |
| **INTRODUCTION** | | | | | |
| Rationale | 6 | Describe the rationale for the review in the context of what is already known | ☒ | ☐ |  |
| Objectives | 7 | Provide an explicit statement of the question(s) the review will address with reference to participants, interventions, comparators, and outcomes (PICO) | ☒ | ☐ |  |
| **METHODS** | | | | | |
| Eligibility criteria | 8 | Specify the study characteristics (e.g., PICO, study design, setting, time frame) and report characteristics (e.g., years considered, language, publication status) to be used as criteria for eligibility for the review | ☒ | ☐ |  |
| Information sources | 9 | Describe all intended information sources (e.g., electronic databases, contact with study authors, trial registers, or other grey literature sources) with planned dates of coverage | ☒ | ☐ |  |
| Search strategy | 10 | Present draft of search strategy to be used for at least one electronic database, including planned limits, such that it could be repeated | ☒ | ☐ |  |
| ***STUDY RECORDS*** | | | | | |
| Data management | 11a | Describe the mechanism(s) that will be used to manage records and data throughout the review | ☒ | ☐ |  |
| Selection process | 11b | State the process that will be used for selecting studies (e.g., two independent reviewers) through each phase of the review (i.e., screening, eligibility, and inclusion in meta-analysis) | ☒ | ☐ |  |
| Data collection process | 11c | Describe planned method of extracting data from reports (e.g., piloting forms, done independently, in duplicate), any processes for obtaining and confirming data from investigators | ☒ | ☐ |  |
| Data items | 12 | List and define all variables for which data will be sought (e.g., PICO items, funding sources), any pre-planned data assumptions and simplifications | ☒ | ☐ |  |
| Outcomes and prioritization | 13 | List and define all outcomes for which data will be sought, including prioritization of main and additional outcomes, with rationale | ☒ | ☐ |  |
| Risk of bias in individual studies | 14 | Describe anticipated methods for assessing risk of bias of individual studies, including whether this will be done at the outcome or study level, or both; state how this information will be used in data synthesis | ☒ | ☐ |  |
| ***DATA*** | | | | | |
| Synthesis | 15a | Describe criteria under which study data will be quantitatively synthesized | ☒ | ☐ |  |
|  | 15b | If data are appropriate for quantitative synthesis, describe planned summary measures, methods of handling data, and methods of combining data from studies, including any planned exploration of consistency (e.g., *I*^2^, Kendall’s tau) | ☒ | ☐ |  |
|  | 15c | Describe any proposed additional analyses (e.g., sensitivity or subgroup analyses, meta-regression) | ☒ | ☐ |  |
|  | 15d | If quantitative synthesis is not appropriate, describe the type of summary planned | ☒ | ☐ |  |
| Meta-bias(es) | 16 | Specify any planned assessment of meta-bias(es) (e.g., publication bias across studies, selective reporting within studies) | ☒ | ☐ |  |
| Confidence in cumulative evidence | 17 | Describe how the strength of the body of evidence will be assessed (e.g., GRADE) | ☐ | ☒ |  |

TABLE 3: Quality assessment of the seven studies included in the meta analysis

|  | **Selection** | | | | **Comparability** | **Outcome** | | | **Score** |
| --- | --- | --- | --- | --- | --- | --- | --- | --- | --- |
| Author | Representativeness of the exposed cohort: | Selection of the non exposed cohort | Ascertainment of exposure | outcome of interest was not present at start of study | Comparability of cohorts on the basis of the design or analysis | Assessment of outcome | Was follow-up long enough for outcomes to occur | Adequacy of follow up of cohorts | SCORE |
| [Honda](https://www.ncbi.nlm.nih.gov/pubmed/?term=Honda%20Y%5BAuthor%5D&cauthor=true&cauthor_uid=27324158) (35) | * | * | * | * | ** | * | * | NS | 8 |
| [Cheng YL](https://www.ncbi.nlm.nih.gov/pubmed/?term=Cheng%20YL%5BAuthor%5D&cauthor=true&cauthor_uid=28649089)(12) | * | * | * | * | ** | * | * | NS | 8 |
| [Shirakabe A](https://www.ncbi.nlm.nih.gov/pubmed/?term=Shirakabe%20A%5BAuthor%5D&cauthor=true&cauthor_uid=28803356)(34) | * | * | * | * | ** | * | * | NS | 8 |
| [Iwakami N](https://www.ncbi.nlm.nih.gov/pubmed/?term=Iwakami%20N%5BAuthor%5D&cauthor=true&cauthor_uid=28041709)(32) | * | * | * | * | ** | * | * | NS | 8 |
| Sze S(36) | ***** | ***** | ***** | ***** | ****** | ND | ***** | NS | **7** |
| Ouchi S(37) | * | * | * | * | ** | * | * | * | 9 |
| Jae Yeong Cho(33) | * | * | * | * | ** | * | * | NS | 8 |

Table 4: Nutrition screening instrument parameters used in review studies.

| PNI | | | | | | | | | | | |
| --- | --- | --- | --- | --- | --- | --- | --- | --- | --- | --- | --- |
| Total score category£ | | Normal: High-PNI | | | | | Moderate: Middle-PNI | | | | Severe: Low-PNI |
| Dysnutritional state PNI cut off [34,36] | | >38 | | | | | 35 to 38 | | | | <35 |
| Dysnutritional state PNI cut off[12] | | >44.8 | | | | | 39.3 to 44.8 | | | | ≤39.3 |
| GNRI | | | | | | | | | | | |
| Total score category | Normal | | Mild | | | | | Moderate | | Severe | |
|  | > 98 | | 98-92 | | | | | 91-82 | | <82 | |
| Dysnutritional state GNRI cut off ¥ |  | | GNRI ≤ 98[36] | | | | | | | | |
|  |  | | | | | | | GNRI < 92[35,37] | | | |
| NRI | | | | | | | | | | | |
|  | Normal | | | | Mild | Moderate | | | Severe | | |
| Total score category[33] | >100 | | | | 95-100 | 89-95 | | | <89 | | |
| CONUT | | | | | | | | | | | |
| Albumin, g/dL (score) | ≥ 3.5 (0) | | | 3.0–3.4 (2) | | | | 2.5–2.9 (4) | | <2.5 (6) | |
| Total lymphocyte count/mL (score) | ≥1600 (0) | | | 1200–1599 (1) | | | | 800–1199 (2) | | <800 (3) | |
| Total cholesterol, mg/dL (score) | ≥180 (0) | | | 140–179 (1) | | | | 100–139 (2) | | <2.5 (3) | |
| Total score category [32,34] | Normal (0–1) | | | Mild (2–4) | | | | Moderate (5–8) | | Severe (9–12) | |
| Dysnutritional state CONUT cut offµ[36] |  | | | | | | | CONUT > 4 | | | |

^£:^ Prognostic nutritional index was used in 3 studies, the total score category in Cheng and Al.'s study was different compared to the two others. PNI= 10 x serum albumin in g/dL+ 0.005 x total lymphocyte count in mm^3^ (Lower = Worse). GNRI= 1.489 x serum albumin in g/dL+ 41.7 (BW in kg/ideal BW*); The ideal body weight (IBW) = 22 x square of height in meters. IBW was calculated by the Devine formula for men (IBW [kg] = 50 kg + 2.3 kg for each inch of height > 5 feet) and the Robinson formula for women (IBW [kg] 48.67 kg + 1.65 kg for each inch of height over 5 feet); NRI= 1.519 x serum albumin in g/dL+41.7x (BW in kg/Ideal BW*); CONUT= using serum albumin, total lymphocytes count and total cholesterol. ^¥^: Geriatric Nutritional Risk Index was used in 4 studies. GNRI cut off value defined by Sze and Al. was lower compared to the 3 others study. ^µ^ Among the 4 studies using CONUT, only Sze and Al. defined a cut-off, the 3 others used the 4 total score category.

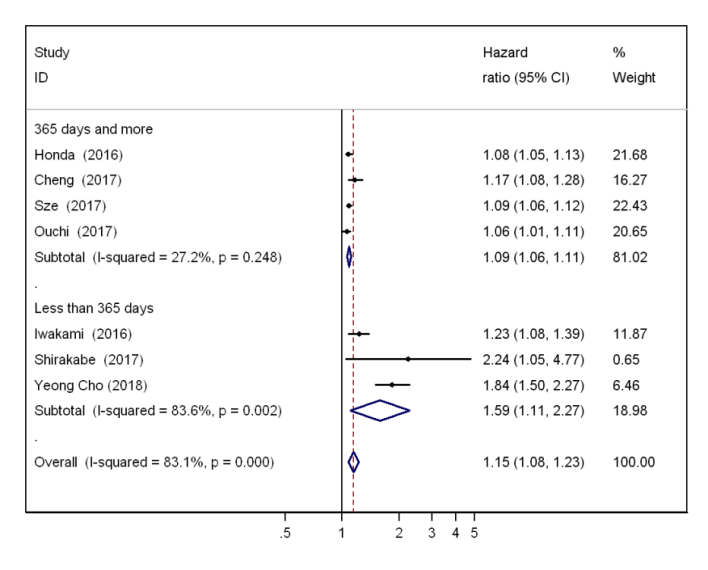
